# A Rapid Review of Advance Care Planning Interventions, Strategies, and Communication Approaches in Dementia Care

**DOI:** 10.1101/2025.08.28.25334659

**Authors:** Tharin Phenwan, Thagoon Kanjanopas, Kanthee Anantapong, Supakorn Sripaew, Jiroj Phalalert, Atiqur Rahman

## Abstract

Advance care planning is essential for aligning future care with the values and preferences of people living with dementia and their families. Challenges from people with dementia’s fluctuating mental capacity, gradual decline and healthcare professional’s limited advance care planning knowledge and skills remain to enable people with dementia to fully engage with the process; tailored interventions are needed.

This rapid review synthesised evidence on advance care planning interventions, communication strategies and health-related outcomes in dementia care. Following Cochrane’s guidance for rapid reviews, we searched CINAHL, Cochrane Central, PubMed, and Web of Science through May 2025. After duplicate removal, title-abstract and full-text screening were conducted in Covidence with dual independent reviewers. Data extraction and quality assessment, using Joanna Briggs Institute tools, employed a single-reviewer approach with verification by second reviewer.

Twenty-five studies from 2015–2025 across 12 countries met inclusion criteria. Included articles were of quantitative designs (n=15), qualitative (n=5) and mixed methods (n=4).

Interventions fell into three categories: video as decision aids; web-based tools; and multicomponent programmes combining education, structured discussions, and documentation support for people with dementia, families and healthcare professionals.

Primary outcomes consistently showed increased advance care planning uptake. Certain secondary outcomes—carer burden, cost of care, carer’s sense of competence, hospitalisation rates, quality-adjusted life year, quality of life of people with dementia, rate of burdensome treatments—demonstrated mixed results. Communication strategies identified included embedding relevant theories such as relational autonomy and shared decision-making frameworks for advance care planning process.

Study quality ranged from poor (n=8) to high (n=6). Common limitations include small sample sizes, unclear randomisation and allocation processes and limited reflexivity in qualitative research. These findings suggest that contextually tailored advance care planning interventions improve uptake but require standardised outcomes and broader cultural adaptation to comprehensively assess impacts on health outcomes.

## Introduction

Advance care planning is an iterative process which individuals articulate, discuss and document their future care preferences with healthcare professionals and trusted proxies to ensure future care aligns with their values and goals when they are unable to make decisions themselves (van der Steen et al., 2024). Advance care planning has been associated with improved quality of life, greater satisfaction with care, reduced conflicts and avoidance of unnecessary treatments (Malhotra et al., 2022; McMahan et al., 2021). Early advance care planning discussion is beneficial and is recommended since it ensures that families, healthcare professionals involved are aware of people with dementia’s preferences of care and can appropriately support their needs (Phenwan, Sixsmith, et al., 2025).

Nevertheless, people with dementia face unique challenges to fully engage with advance care planning. Their symptoms such as anxiety, forgetfulness and fluctuating mental capacity complicate timely discussions, and families may struggle to interpret the person’s preferences, particularly during advanced stages, that are reflective to their changing needs (Malhotra et al., 2022; McMahan et al., 2021; Phenwan, Sixsmith, et al., 2025). Healthcare professionals also report limited training and reluctance to initiate advance care planning in dementia care, further reducing uptake (Malhotra et al., 2022).

To address these challenges, customised advance care planning interventions—including structured conversations, decision aids and family-centred programmes—have been proposed to support meaningful dialogue and documentation (Wendrich-van Dael et al., 2020). However, previous reviews on advance care planning in dementia were limited by date or design focus— either covering literature only up to 2018 (Wendrich-van Dael et al., 2020) or 2020 (McMahan et al., 2021), or including exclusively quantitative trials (Kelly et al., 2019; McMahan et al., 2021)— thereby omitting qualitative insights into contextual facilitators and barriers that influence the uptake of advance care planning. A comprehensive, up-to-date synthesis that captures both intervention effectiveness and the mechanisms underlying advance care planning uptake is needed.

This rapid review aims to i) synthesise existing evidence in relation to advance care planning interventions and communication strategies with and for people with dementia and ii) evaluate the effectiveness of these interventions and communication strategies in increasing the uptake of advance care planning and improving related health outcomes. We included the communication strategies given they were inherent to advance care planning thus can influence its uptake and effectiveness.

The review questions are:

*What interventions or communication strategies have been implemented to promote the uptake of advance care planning/advance directive with and for people with dementia?*

*Which health-related outcomes have been measured?*

*How effective are these interventions or strategies in enhancing advance care planning uptake and health outcomes?*

## Methods

A rapid review approach based on Cochrane’s Updated guidance on methods used in Cochrane rapid reviews of effectiveness was undertaken (Garritty et al., 2024). The approach was chosen to balance the time constraints and the robustness of the process while maintain transparency of the review process, using their recommendations. The review protocol was registered with the International Prospective Register of Systematic Reviews database (CRD420251034120).

## Knowledge user engagement

Five of the research team have been appointed as a sub-team for the revised Thai dementia national guideline working group and were responsible for creating recommendations around advance care planning interventions with and for people with dementia.

The review questions and the chosen rapid review approach had been consulted with, co-developed and mutually agreed with stakeholders, ensuring that we could deliver recommendations in a timely manner and contextual relevance. Initial findings from this review have been reported to the stakeholders and the panel of experts in July 2025 and will be used as a part of the revised Thai national guideline.

### Eligibility criteria

Studies were included based on the inclusion and exclusion criteria (see Table 1). The PICO framework was used to formulate and frame the criteria and review questions.

**Table 1.** Inclusion and exclusion criteria.

|  | Inclusion criteria | Exclusion criteria |
| --- | --- | --- |
| Population | Articles that discussed:<br>-adults who have been diagnosed with dementia<br>-family carers of people with dementia involved in advance care planning/advance directive discussion<br>-healthcare professionals involved in advance care planning/advance directive discussion | -not related to people with dementia, family carers of people with dementia, healthcare professionals caring for people with dementia<br>-paediatrics |
| Intervention(s) or exposure(s) | -any intervention or communication strategy aimed at increasing advance care planning/advance directive uptake (e.g., structured conversations, decision aids, family-centred approach, digital tools) | -interventions or communication strategy unrelated to advance care planning/advance directive with and for people with dementia |
| Comparison with usual care | any contexts of care (e.g., community, primary care, acute hospitals, long-term care facilities) |  |
| Outcomes | *Primary outcome:*<br>- advance care planning/advance directive uptake or completion rate<br>*Secondary outcomes:*<br>-quality of life<br>-decisional conflict<br>-healthcare utilisation metrics<br>-quality of communication<br>-concordant with care preferences<br>-discordant of care<br>-hospitalisation rate<br>-carer burden<br>-place of death | -No outcomes measured nor mentioned |
|  | -peer-reviewed articles<br>-published after 2015<br>-published in English<br>-Any region | -non-peer reviewed articles<br>-theoretical, ethical, legal discussion articles<br>-reviews |

### Search strategy

Five team members, supported by an academic librarian, searched CINAHL, Cochrane Central, PubMed and Web of Sciences through May 2025. A combination of keywords, Boolean operators, and truncations was applied to maximise retrieval of relevant studies (see Supplementary file 1).

### Screening and selection

The search results were uploaded into Covidence, a review management software, for semi-automated duplicate removal. Duplications were further removed by manual checking.

Initially, six reviewers performed 25 title screening pilot together against the inclusion and exclusion criteria and discussed any ambiguities. We then used the dual screening process. Six reviewers were randomly assigned independently to screen the articles, minimising selection bias. The lead reviewer was responsible for final decision whether to include or exclude the articles in case of discrepancies. Titles with unclear or absent abstract were included for subsequent full-text review.

Similar process was applied during full-text review, staring with pilot screening against the inclusion and exclusion criteria and the dual screening process of randomly assigned articles with the lead review solving discrepancies.

### Data extraction

We undertook a single-reviewer extraction approach with a second reviewer verifying the accuracy of extractions. A pilot extraction file was created in Covidence. The lead reviewer then extracted relevant data to the pilot extraction file; the process was iterative, ensuring the review questions were fully answered. Once the extraction file has been finalised, the lead reviewer continued to extract the remaining data until completion. Remaining reviewers then verified the completeness of the extracted data.

The extracted data includes:

*Titles, authors, year, country, study design, context of the study, target audience of interventions, persons delivering interventions and process, quantitative primary outcomes, qualitative primary outcomes, quantitative secondary outcomes, qualitative secondary outcomes*

Study authors were not contacted for clarification or obtaining information of missing data to ensure a timely completion of the review.

### Quality assessment

All included studies were appraised for their quality and risk of bias using relevant Joanna Briggs Institute critical appraisal tools (Aromataris et al., 2024). This process was performed by the lead reviewer. Two reviewers were then randomly assigned to verify the judgement to ensure both methodological robustness and timely completion of the review. No articles were excluded based on quality assessment given we want to comprehensively capture existing literature.

### Data synthesis

Due to the anticipated heterogeneity in study designs and outcomes, findings were narratively synthesised. The included studies were categorised with their shared characteristics and findings were related back to the review questions.

A meta-analysis was not conducted. The Preferred Reporting Items for Systematic Reviews and Meta-Analyses statement (Stevens et al., 2025) was used to indicate the list of key reporting items for the rapid review process (see Supplementary file 2).

## Analysis

The included databases were searched up to May 2025. A total of 1,350 records were identified through database searches. After removing 855 duplicates via Covidence and manual checking, 495 titles and abstracts were screened. 74 articles were included for full-text assessment. Of which, six were not available; 49 articles were subsequently excluded, leaving 25 included articles for the review (see Figure 1.)

**Figure 1.**
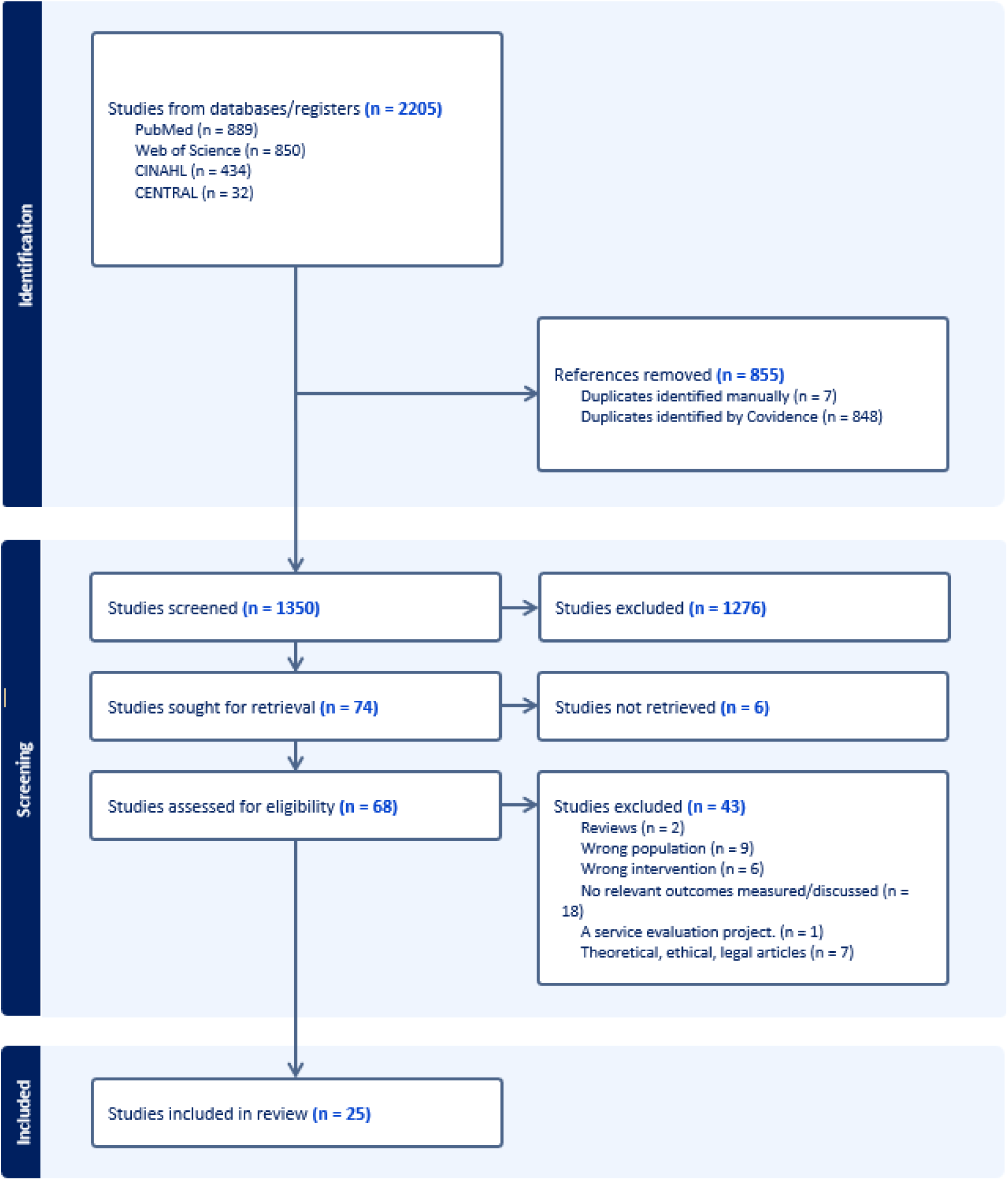
PRISMA flowchart

### Study characteristics

The included 25 studies were published between 2015-2025 and spanned 12 countries: USA (n=7) (Bonner et al., 2021; Cohen et al., 2019; Cotter et al., 2019; Jennings et al., 2019; Kistler et al., 2021; Mellinger et al., 2023; Mitchell et al., 2018), Canada (n=3) (Ménard et al., 2025; Sussman et al., 2022; Vellani et al., 2023), UK (n=3) (Brazil et al., 2018; Davies et al., 2021; Moore et al., 2017), Belgium (n=3) (Dupont et al., 2024;

Monnet et al., 2024; Wils et al., 2017), the Netherlands (n=2) (Tilburgs et al., 2020; van Soest-Poortvliet et al., 2015), Singapore (n=1) (Grp et al., 2021), Switzerland (n=1) (Bosisio et al., 2021), Spain (n=1) (Sarabia-Cobo et al., 2016), Hongkong, China (n=1) (Yeung et al., 2023), Taiwan, China (n=2) (Huang et al., 2022; Huang et al., 2020). One study was conducted in 6 countries (Canada, Czech Republic, Ireland, Italy, the Netherlands and the UK) (Bavelaar et al., 2023), indicating a universal interest of the topic.

Eleven studies were conducted in community settings (Bonner et al., 2021; Davies et al., 2021; Dupont et al., 2024; Grp et al., 2021; Kistler et al., 2021; Ménard et al., 2025; Monnet et al., 2024; Sussman et al., 2022; Tilburgs et al., 2020; Vellani et al., 2023; Yeung et al., 2023). Eight focussed in long-term care facilities (Bavelaar et al., 2023; Brazil et al., 2018; Cohen et al., 2019; Mitchell et al., 2018; Moore et al., 2017; Sarabia-Cobo et al., 2016; van Soest-Poortvliet et al., 2015; Wils et al., 2017), followed by four studies conducted in acute hospital settings (Cotter et al., 2019; Huang et al., 2022; Huang et al., 2020; Jennings et al., 2019). Only two studies were conducted in multiple settings (Bosisio et al., 2021; Mellinger et al., 2023).

Regarding study designs, 16 studies were quantitative designs. Of which, seven used a quasi-experimental design (Bosisio et al., 2021; Cotter et al., 2019; Huang et al., 2022; Huang et al., 2020; Vellani et al., 2023; Wils et al., 2017; Yeung et al., 2023). Seven were randomised controlled trials (Bavelaar et al., 2023; Bonner et al., 2021; Brazil et al., 2018; Cohen et al., 2019; Mellinger et al., 2023; Mitchell et al., 2018; Tilburgs et al., 2020) and one study used a prospective cohort design (Huang et al., 2022). Five studies utilised qualitative methods (Davies et al., 2021; Grp et al., 2021; Ménard et al., 2025;

Sarabia-Cobo et al., 2016; van Soest-Poortvliet et al., 2015) and four were mixed methods studies (Dupont et al., 2024; Monnet et al., 2024; Moore et al., 2017; Sussman et al., 2022) (See Table 2).

**Table 2.**
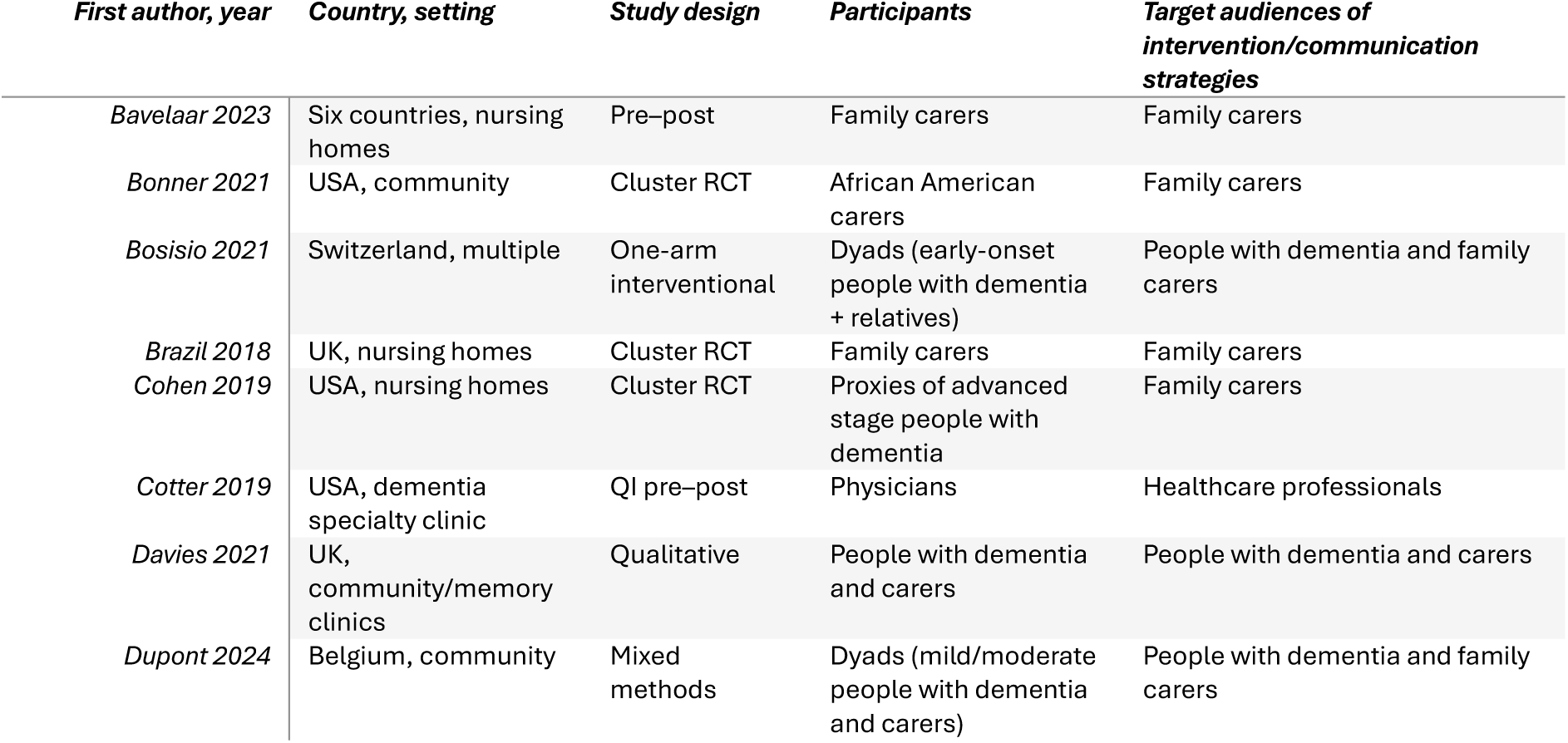
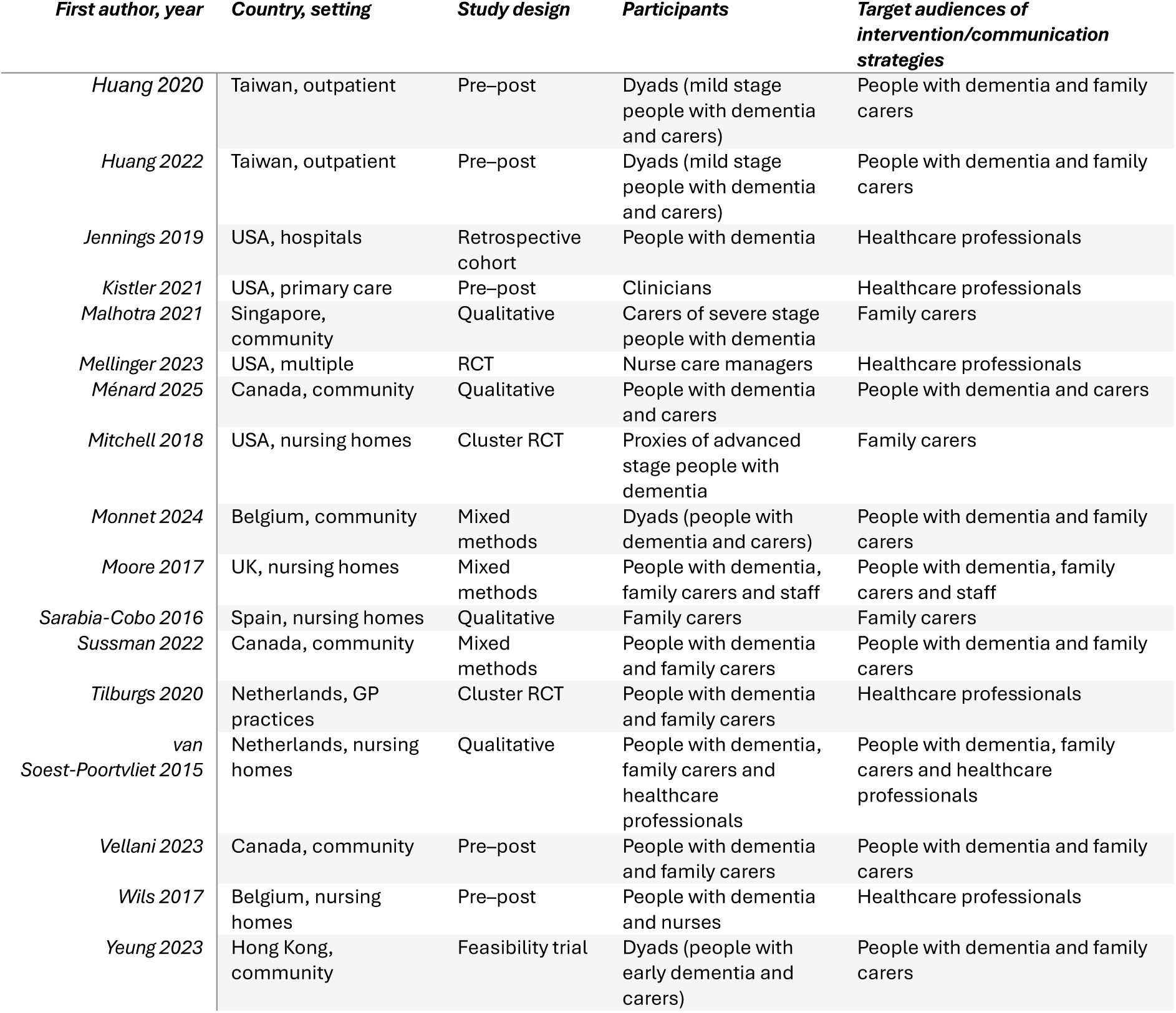
Characteristics of included studies.

No study focused exclusively on people with dementia. This is expected given their symptoms of fluctuating mental capacity and gradual decline hence the interventions tend to tailor for people with dementia along with other stakeholders involved over the advance care planning process.

Ten articles focused on people with dementia and carers as dyads (Bosisio et al., 2021; Davies et al., 2021; Dupont et al., 2024; Huang et al., 2022; Huang et al., 2020; Ménard et al., 2025; Monnet et al., 2024; Sussman et al., 2022; Vellani et al., 2023; Yeung et al., 2023) while seven focused on family carers (Bavelaar et al., 2023; Bonner et al., 2021; Brazil et al., 2018; Cohen et al., 2019; Grp et al., 2021; Mitchell et al., 2018; Sarabia-Cobo et al., 2016). Six articles targeted healthcare professionals delivering care for people with dementia (Cotter et al., 2019; Jennings et al., 2019; Malhotra et al., 2022; Mellinger et al., 2023; Tilburgs et al., 2020; Wils et al., 2017) and two aimed at both family carers and healthcare professionals (Moore et al., 2017; van Soest-Poortvliet et al., 2015).

Three articles included people with dementia from multiple stages (Bosisio et al., 2021; Dupont et al., 2024; Monnet et al., 2024). The more inclusion of people with dementia from various clinical stages is reassuring given their capacity to consent and participate is research do not correlate with their clinical staging. The inclusive research designs then contribute to assert people with dementia’s human rights to participate in research without being prematurely excluded (Phenwan, 2021).

### Quality appraisal of the included studies

Studies were rated as: medium quality (n= 11) (Bonner et al., 2021; Brazil et al., 2018; Cohen et al., 2019; Davies et al., 2021; Grp et al., 2021; Huang et al., 2020; Jennings et al., 2019; Mitchell et al., 2018; Tilburgs et al., 2020; Wils et al., 2017; Yeung et al., 2023); poor quality (n=8) (Bosisio et al., 2021; Brazil et al., 2018; Cotter et al., 2019; Huang et al., 2022; Kistler et al., 2021; Mellinger et al., 2023; Sarabia-Cobo et al., 2016; Vellani et al., 2023) and high quality (n= 6) (Bavelaar et al., 2023; Dupont et al., 2024; Ménard et al., 2025; Monnet et al., 2024; Sussman et al., 2022; van Soest-Poortvliet et al., 2015) (see Supplementary file 3).

For quantitative and mixed-methods studies, common limitations included small sample sizes that reduced statistical power, unclear or biased sampling procedures and insufficient use of statistical controls to account for confounding variables. Many studies employed cross-sectional or short-duration designs, which limited causal inference and temporality. The absence of control groups—often due to ethical or practical constraints—further weakened the ability to attribute observed effects to the intervention.

For qualitative study, the lack of details regarding researcher’s positionalities or identities that influence the study design and data analysis process was the primary reason for poor quality studies. Researchers’ reflexivity also tends to be unstated or formulaically described with limited nuances from the studies.

### Advance Care Planning interventions with and for people with dementia

Three types of interventions have been identified (see Table 3):

1. Using videos as decision aids
2. Interactive websites that provide dementia and advance care planning information and tools.
3. Multicomponent interventions tailored to stakeholders. The intervention modalities ranged from: i) education sessions for GPs and nurses, physicians at dementia specialty clinic, nurses, family carers, people with dementia and family carers and
4. educational sessions plus other activities such as facilitated structured discussion, advance care planning documentation support, coordination of care between care teams and advance care planning booklets.

**Table 3.**
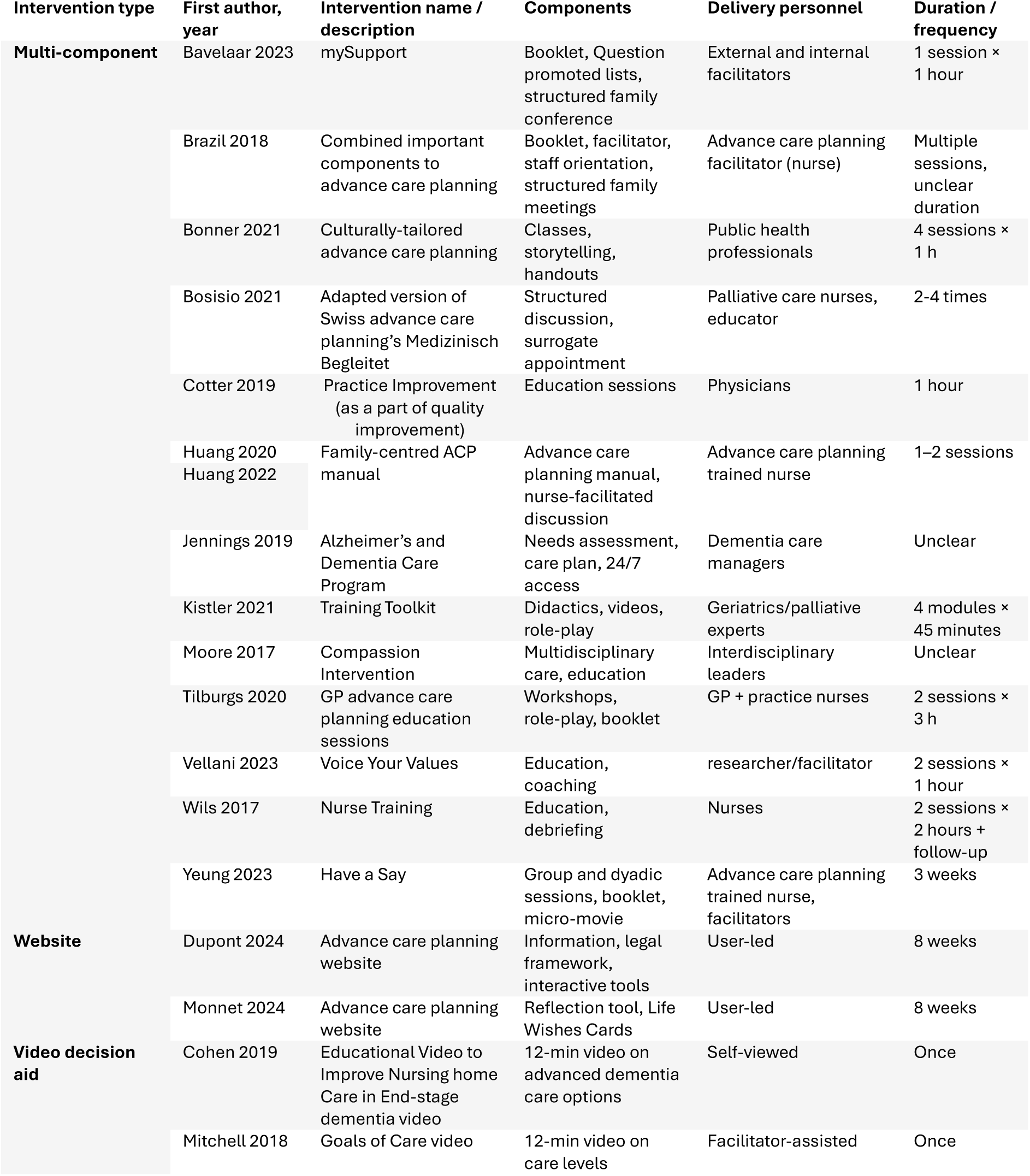
Summary of ACP interventions by type.

### Proposed communication strategies

This section focuses on the five studies that did not offer explicit advance care planning interventions but offered theoretical or communication frameworks to promote its uptake.

### Addressing the lack of dementia knowledge in families

Sarabia-Cobo et al. (2015) suggested that it is essential to provide more dementia knowledge for families given the lack of knowledge, particularly of dementia progression, contributes to their inability to conceptualise people with dementia’s final stages of life and necessary choices they need to consider along the way (Sarabia-Cobo et al., 2016). The authors proposed websites as both tools and strategies to improve concordance of care:

> *‘[participants who showed the highest degree of agreement with advance directives and decisions] had searched for information through the Alzheimer’s Association and other [websites for] support…*’ (Sarabia-Cobo et al., 2016), p. e9This proposal resonated well with the web-based interventions identified in this review.

### Using relevant frameworks or robust theories for advance care planning

Menard et al. (2025) suggested using a relational autonomy as a framework to discuss advance care planning with and for people with dementia and their carers. The discussions need to be comprehensive and included practical issues such as advice and support around home modifications (Ménard et al., 2025). This strategy is based on an assertion that as the health of people with dementia declines, their autonomy and carers will be intertwined thus the upcoming decisions are influenced by this interdependence.

This suggestion is similar to what Davies et al. (2021). They used The Interprofessional Shared Decision Making model as a conceptual model of decision-making in dementia end of life care (Davies et al., 2021). Their comprehensive model aims to address previous existing advance care planning models that do not holistically capture people with dementia and families’ changing needs. They also intend to mitigate unclear expectations from both people with dementia and families since, without a clearer guidance, both *‘appeared to not fully understand what the decisions [around advance care planning] meant and how they would help them in the future.’* (Davies et al., 2021) p7.

Similarly, Malhotra et al. (2022) proposed addressing the discordance over goals of care between family carers of community-dwelling older adults with severe dementia and the extended families in Singapore using the Respecting Choices Model for advance care planning. Due to the collectivist cultural context, carers may feel coerced to choose life-prolonging treatments even though such treatment is not consistent with the expressed treatment preferences discussed with and for people with dementia. The authors then proposed strategies to mitigate this discordance by a more comprehensive discussions including both overall end-of-life care goal and common clinical scenarios for people with dementia, ensuring that caregivers’ have a better dementia knowledge and understanding of the consequences of the treatment preferences (Malhotra et al., 2022).

Finally, van Soest-Poortvliet et al. (2015) (van Soest-Poortvliet et al., 2015) suggested two strategies to facilitate the advance care planning process: proactive discussion and reactive discussion. Being proactive means physicians should actively initiate the discussion around end-of-life care before expanding beyond that. They can then discuss possible treatments by outlining various scenarios that are relevant and co-created tailored-made goals with and for people with dementia and families over time. Conversely, being reactive means physicians can utilise the ‘wait and see’ approach and use each opportunity as a leverage to start discussing advance care planning e.g., a change in people with dementia’s condition (van Soest-Poortvliet et al., 2015). These two-pronged strategies aim to balance the necessity of timely discussion while being pragmatic and not ‘*presenting an entire trajectory about something that may or may not happen’, p984* See table 4.

**Table 4.**
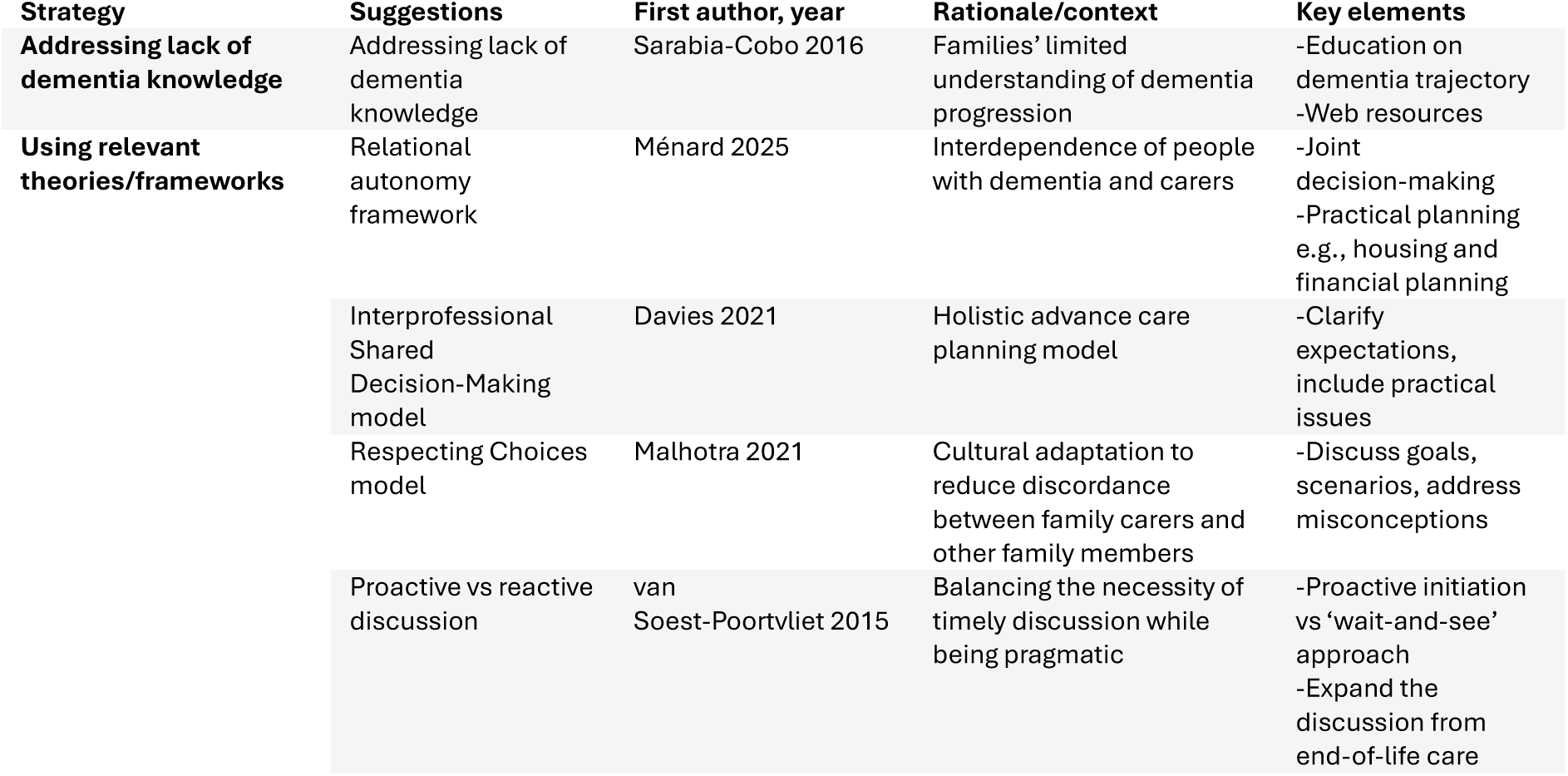
Communication strategies identified.

### Reported health outcomes

#### Primary outcomes

Eight articles reported the increased uptake of advance care planning (Bavelaar et al., 2023; Bonner et al., 2021; Dupont et al., 2024; Huang et al., 2022; Huang et al., 2020; Mellinger et al., 2023; Sussman et al., 2022; Wils et al., 2017). Nine focused on the increased documentation of advance directive or end-of-life care medical decisions (Bosisio et al., 2021; Brazil et al., 2018; Cotter et al., 2019; Huang et al., 2022; Jennings et al., 2019; Kistler et al., 2021; Mitchell et al., 2018; Tilburgs et al., 2020; Wils et al., 2017) or a combination of both. These vacillating outcomes are expected given the evolving focus and definition of advance care planning over time.

The terminologies used in the articles were largely variable, making it problematic since each term has a different focus within the concept of advance care planning. The terms used are:

Advance directive, Advance care preference, Advance decision to refuse treatment, Do not attempt cardiopulmonary resuscitation, Do not intubate, Do not resuscitate, Health care power of attorney, Living will, Medical orders for life-sustaining treatment, Power of attorney, Physician orders for life-sustaining treatment.

In general, older articles tend to focus on end-of-life medical decisions while the more recent articles are much broader and include non-medical discussions and decisions, reflecting the shifting focus of advance care planning.

#### Secondary outcomes

The secondary outcomes were measured by validated instruments. However, the captured outcomes were vastly variable, making it challenging to compare the effectiveness of the identified advance care planning interventions given most included studies focused on different outcomes. The identified outcomes include:

Advance care planning engagement, Any hospitalisation record, Attitudes toward advance care planning, Carers’ burden via the Zarit Burden Scale, Carer’s sense of competence, Cost of care, Concordance of care between people with dementia and carers, Decisional Conflict,

Decisional Autonomy, Family Perceptions of Care, Knowledge of advance care planning, Level of shared-decision making, Numbers of surrogated being appointed for people with dementia, Numbers of acute care events or emergency visit of people with dementia, Numbers of ICU stay, Place of death, Quality-adjusted life year, Quality of life of people with dementia, The rate of burdensome treatments, Satisfaction with care at end-of-life in dementia.

Table 5 summarises the identified primary and secondary outcomes.

**Table 5.** Primary and secondary outcomes.

| Outcome category | Measure(s) | Studies reporting | Direction of effect |
| --- | --- | --- | --- |
| **Primary outcomes** |  |  |  |
| Advance care planning uptake | % with advance care planning/advance directive documented | 17/25 studies | ↑ |
| **Secondary outcomes** |  |  |  |
| Concordance | % agreement, kappa | Bonner et al., 2021<br>Bosisio et al., 2021<br>Cohen et al., 2019<br>Huang et al., 2020<br>Huang et al., 2022<br>Yeung et al., 2023 | ↑ |
| Carer's sense of competence | Dutch version of the Sense of Competence Questionnaire | Tilburgs et al., 2020 | No statistically significant in two groups |
| Cost of care | Dutch version of the Resource Utilization in Dementia questionnaire | Tilburgs et al., 2020 | No statistically significant in two groups |
| Decisional Autonomy | Decisional Autonomy Scale | Bosisio et al., 2021 | ↑ |
| Decisional conflict | Decisional conflict Scale | Bavelaar 2023<br>Brazil 2018<br>Huang 2020 | ↓ |
| Attitudes | Advance care planning attitudes scale | Bonner et al., 2021<br>Huang et al., 2020 | ↑ |
| Knowledge | Advance care planning knowledge scale | Huang et al., 2020<br>Bonner et al., 2021 | ↑ |
| Engagement | Advance care planning engagement scale | Bonner et al., 2021<br>Dupont et al., 2024<br>Sussman et al., 2022<br>Vellani et al., 2023<br>Yeung et al., 2023 | ↑ |
| Family perceptions of care | Family perceptions of care scale | Bavelaar et al., 2023<br>Brazil et al., 2018 | ↑ |
| Carer burden | Zarit Burden Scale | Bosisio et al., 202<br>Yeung et al., 2023 | Mixed |
| Hospitalisation | Hospitalisation rate | Jennings et al., 2019<br>Bavelaar et al., 2023 | Mixed |
| Level of shared-decision making | CollaboRATE questionnaire | Tilburgs et al., 2020 | No statistically significant in two groups |
| Numbers of surrogated being appointed for people with dementia | Numbers of surrogated appointed post-intervention | Bosisio et al., 2021<br>Kistler et al., 2021 | ↑ |
| Numbers of acute care events or emergency visits of people with dementia | Numbers of events | Jenning et al., 2019 | ↓ |
| Numbers of ICU stay | Numbers of events | Jenning et al., 2019 | No statistically significant in two groups |
| Home death | Numbers of home death | Jenning et al., 2019 | ↑ |
| Quality-adjusted life year | The EuroQOL 5D utility scores with survival | Tilburgs et al., 2020 | No statistically significant in two groups |
| Quality of life | Dementia quality of Life questionnaire | Tilburgs et al., 2020 | No statistically significant in two groups |
| Rate of burdensome treatments | % of any hospital transfers, feeding tube insertions, or parenteral therapy | Mitchell et al., 2018 | No statistically significant in two groups |
| Satisfaction with care at end-of-life in dementia | Satisfaction with care at end-of-life in dementia scale | Moore et al., 2017 | ↑ |

### Intervention effects over advance care planning uptake

#### Primary outcomes

The identified advance care planning interventions has demonstrated the increased uptake, discussion and documentation (Bavelaar et al., 2023; Bonner et al., 2021; Bosisio et al., 2021; Brazil et al., 2018; Cotter et al., 2019; Dupont et al., 2024; Huang et al., 2022; Jennings et al., 2019; Kistler et al., 2021; Mellinger et al., 2023; Mitchell et al., 2018; Sussman et al., 2022; Tilburgs et al., 2020; Wils et al., 2017). For instance, Bavelaar et al. (2023)’s study of nursing homes in six countries found that their intervention, mySupport, had statistically significant improved the numbers of advance decision to refuse treatment (24% versus 32% in pre- and post-intervention period, respectively with p = 0.006 (Bavelaar et al., 2023). This result is similar to Mitchell et al. (2018)’s randomised trial that used a video as a decision support tool as an intervention for family carers. In their study, participants in the control arm from their study with advance directives in place had forgone tube feeding at all assessments period with the adjust odds ratio of 2.32 and 95% confidence interval of 1.38-3.91 at month 12 (Mitchell et al., 2018).

Qualitative feedback from the studies indicated that the interventions has facilitated advance care planning initiation. However, participants were often unclear about next steps particularly for a user-led intervention without facilitators such as a website:

*’They [both people with dementia and carers] found the [incorporated advance care planning] tools [on website] a good way to initiate conversations but were unsure about what the next step should be…’ (Monnet et al., 2024), p.6*

This insight relates to the identified strategies earlier since both people with dementia and their families tend to need a clearer guidance to facilitate their expectations (Davies et al., 2021; Ménard et al., 2025; Sarabia-Cobo et al., 2016).

#### Secondary outcomes

We found mixed findings regarding the effects of advance care planning interventions over the identified secondary outcomes.

### Positive secondary outcomes Advance care planning engagement

The identified interventions seem to improve advance care planning engagement for their target audiences (people with dementia, carers or both) (Bonner et al., 2021; Dupont et al., 2024; Sussman et al., 2022; Vellani et al., 2023; Yeung et al., 2023). Yeung’s feasibility trial found that people with dementia in the intervention group reported a significantly greater improvement, with large effect sizes, in self-efficacy for advance care planning at T3 (p = 0.030; d = 0.98) and readiness for advance care planning at T1 (p = 0.006; d = 1.23) but not at all time points (Yeung et al., 2023).

Sussman et al. (2022) echoed this result as their participants’ who used the advance care planning booklet had an average advance care planning engagement score of 3.94 (SD = 0.67), indicating the moderate level of engagement (Sussman et al., 2022). Qualitative insights from both people with dementia and carers suggested the workbook helpful because it could *‘take the stress away’ and ensure that ‘there’s no guessing [regarding advance care planning]’*.

However, the identified studies were primarily feasibility or pilot studies; they tend to have small sample size thus limiting their generalisibility.

### Concordance of care

Similarly, these interventions improved the concordance of *care (Bonner et al., 2021; Bosisio et al., 2021; Cohen et al., 2019; Huang et al., 2022; Huang et al., 2020; Yeung et al., 2023).* In Cohen et. al (2019)’s cluster randomized clinical trial, when comfort care was preferred, concordance was higher in the intervention group versus control group (10.8% vs. 2.5%; adjusted odds ratio, 2.48; 95% confidence interval, 1.01-6.09). However, only 7% of comfort care option was deemed concordant, indicating a small % of overall preferences of care (Cohen et al., 2019).

Huang et. al (2022) found that their culturally-tailored advance care plan intervention has demonstrated an increased consistency between people with dementia and carers for end-of-life decision post-intervention (ranging from 72.7–77.3%) as well as kappa values (ranging from 0.31–0.37). The kappa p-values indicated significant increases post-intervention in consistency between people with dementia and carers’ preferences for cardiopulmonary resuscitation, the use of artificial respirator, and feeding tube (all p < .05). However, the McNemar test indicated no statistically significant difference in consistency (Huang et al., 2022).

### Decisional conflict and decisional autonomy

The identified interventions helped reducing decisional conflict for carers (Bavelaar et al., 2023; Brazil et al., 2018; Huang et al., 2020) and improved their decisional autonomy (Bosisio et al., 2021). Bavelaar et al. (2023) reported significant improvements in the subscales of feeling informed, clarity of values, and decision-making ability, with adjusted mean differences (95% CI) of –13.8 (–25.4 to –2.1), –12.1 (–20.7 to –3.4), and –8.2 (–15.2 to –1.3), respectively. This positive outcome is similar to Huang et. al (2020) which demonstrated a statistically significant lower decisional conflict for end-of-life care on people with dementia (49.88 → 35.11, t = 4.76, *p* < .001, effect size = −0.8) and carers (37.34 → 27.70, t = 3.89, *p* < .001, effect size = −0.6), indicating the effectiveness of these interventions.

Bosiosio et al. (2021)’s pilot study’s multiple components intervention demonstrated the changed decisional autonomy among family carers with the changed mean score of 5.83 at baseline to 8.16 at the end of study, indicating an improved perceived autonomy. However, participants reported a higher Zarit Burden Scale from the mean score of 28.28 at baseline to 31.83 at the end of study. One explanation might be from carers’ increasing involvement over advance care planning as a part of the intervention hence the reported higher burden.

### Family perceptions of care

The interventions improved family perceptions of care (Bavelaar et al., 2023; Brazil et al., 2018). Bavelaar et al. (2023)’s participants had reported the change in the Family Perception of Care Scale specifically in the Family Support (adjusted mean difference = 5.2; 95% CI: 3.2 to 7.2) and Communication (3.3; 95% CI: 1.8 to 4.7) domains (Bavelaar et al., 2023).

### Home death, acute events admission and emergency visits

The presence of Physician orders for life-sustaining treatment has an inverse effect for home death (Jennings et al., 2019). That is, in Jenning’s study, participants who completed their Physician orders for life-sustaining treatment (N=129/70%) has statistically significant likelihood not to die at home compared to those without the document in place (N=82/59%), p.0.04). They reported fewer acute care events or emergency visit between those without the Physician orders for life-sustaining treatment (N=54/29%), compared to those with one (N=94/51%, p 0.03) as well as the hospitalisation rate (Jennings et al., 2019). There is no statically significant between the numbers of ICU admission in two groups.

### Attitudes and knowledge of advance care planning

The interventions increased both the positive attitudes and knowledge of advance care planning with their target audience (Bonner et al., 2021; Huang et al., 2020) as well as reducing negative attitudes (Huang et al., 2020). Huang (2020) reported statistically significant higher knowledge of dementia and advance care planning on people with dementia (t = −2.79, *p* = .008, effect size = 0.5; and t = −4.10, *p* < .001, effect size = 0.6, respectively) and carers (t = −4.77, *p* < .001, effect size = 0.8; and t = −8.22, *p* < .001, effect size = 1.2, respectively). They also found statistically significant lower negative attitudes toward advance care planning on carers (24.27 → 20.65, t = 3.73, *p* = .001, effect size = −0.7).

Satisfaction with care also improved from these interventions (Moore et al., 2017). Moore et al. (2017)’s naturalistic feasibility study in nursing homes showed an improved satisfaction with care in end-of-life in dementia care dementia scale. Participants reported the median (IQR) in their cohort (n=52) as 30 with an IQR between 29-33) and nursing homes in cluster two n=6) reported the score of 34 with an IQR between 28-39.

#### Unclear outcomes

Several secondary outcomes showed ambiguous results. Tilburg’s Cluster Randomized Controlled Trial of advance care planning education sessions with Dutch general practitioners reported no statistically significant between people with dementia’s quality of life, their experienced level of shared decision making and the carers’ sense of competence in both groups. The health care costs and people with dementia’s quality-adjusted life year also did not differ between groups (Tilburgs et al., 2020).

Mitchel et al. (2018)’s clustered randomised trial showed that the rate of burdensome treatments per 1000 resident-days did not differ significantly between arms (intervention arm, mean [SD], 1.23 [2.31]; control arm, 1.42 [5.03]; p =0.32) (Mitchell et al., 2018). The proportion of carers preferring comfort care also did not differ between arms at baseline or at any follow-up interview. These results call into question of the efficacy of the interventions.

Moreover, contrasting to findings from Jenning’s study, Bavelaar et al. (2023) found no significant impact for hospitalization rate of nursing home residents with dementia (Bavelaar et al., 2023). This might be explained from the different characteristics of participants in these two studies as well as the context of the studies (six European countries vs USA) (see Table 6).

**Table 6.** Effectiveness of interventions across included studies.

| First author, year | Primary outcome effects | Secondary outcome effects | Qualitative findings/notes |
| --- | --- | --- | --- |
| Bavelaar 2023 | ↑ Advance decision to refuse treatment 24% → 32%, $p = 0.006$ | ↑ Family Perceptions of Care—Family Support (adjusted mean difference 5.2; 95% confidence interval 3.2–7.2) and Communication (adjusted mean difference 3.3; 95% confidence interval 1.8–4.7)<br>↓ Decisional Conflict (feeling informed, clarity of values, decision-making abilities) ($\Delta -13.8$; 95% CI -25.4 to -2.1; -12.1 (-20.7 to -3.4), and -8.2 (-15.2 to -1.3)<br>no significant impact on hospitalization | |
| Bonner 2021 | | ↑ advance care planning engagement self-efficacy at week 20 (0.70 vs 0.26, $p < 0.0001$) | Culturally tailored, church-based classes; positive effect on confidence rather than completion |
| Bosisio 2021 | ↑ Presence of advance directive 18% → 90% | ↑ Concordance 83% → 100%;<br>↑ Decisional Autonomy 5.83 → 8.16;<br>↑ Surrogate appointed 36% → 81%<br>↑ Zarit Burden 28.28 → 31.83 | Multi-visit facilitation early on; trade-off with caregiver burden noted |
| Brazil 2018 | Completed Do Not Resuscitate form: 42% vs 51%, $p = 0.18$ (not significant) | ↓ Decisional Conflict ($\Delta -10.5$; 95% CI -16.4 to -4.7; $p = 0.001$)<br>↑ Family Perceptions of Care ($\Delta 8.6$; 95% confidence interval 2.3–14.8; $p = 0.001$) | |
| Cohen 2019 |  | ↑ Concordance when comfort care preferred: 10.8% vs 2.5% (adjusted odds ratio 2.48; 95% CI 1.01–6.09) |  |
| Cotter 2019 | ↑ Presence of advance directive 13.6% → 19.7%, $p = 0.045$; ↑ Medical Orders for Life-Sustaining Treatment 11% → 19%, $p = 0.006$ | | |
| Dupont 2024 | ↑ advance care planning discussion: future care 69% (36/52); end-of-life wishes 46% (24/52) | ↑ Readiness—people with dementia 38% → 71%; Carers 68% → 81% | Carer: '...we have mapped it [advance care planning] out so now we can discuss it with someone else more easily.'<br>People with dementia: 'You should not wait to discuss these topics [ACP] until there is a care emergency [...]. I'll try to discuss this with the people who care about me.' |
| Huang 2020 | | ↓ Decisional conflict—people with dementia 49.88 → 35.11 ($t = 4.76$, $p < .001$, $d = -0.8$); carers 37.34 → 27.70 ($t = 3.89$, $p < .001$, $d = -0.6$).<br>↑ Knowledge (people with dementia and carers, all $p \leq .008$)<br>↓ negative attitudes (carers 24.27 → 20.65, $p = .001$, $d = -0.7$) | Nurse-facilitated manual; family-centred discussions |
| Huang 2022 | ↑ Documentation: advance directive for palliative care ($t = -2.05$, $p = .046$); health care agent ($t = -2.58$, $p = .013$); family carers as proxy signer for Do Not Resuscitate form ($t = -2.55$, $p = .015$) | ↑ Consistency people with dementia-carer (72.7%–77.3% at T1); ↑ Kappa 0.31–0.37 with $p < .05$<br>McNemar tests not significant for change | |
| Jennings 2019 | High overall documentation (Do Not Attempt Cardiopulmonary Resuscitation 64%; Physician Orders | Physician Orders for Life-Sustaining Treatment group vs no Physician Orders for Life-Sustaining Treatment group: |  |
| | for Life-Sustaining Treatment 57%; proxy 100%; goals-of-care 99.7%) | ↑ Emergency/acute events 51% vs 39% ($p = .03$)<br>↑ hospitalisation 43% vs 31% ($p = .04$)<br>↑ home death 70% vs 59% ($p = .04$)<br>ICU stay not significant | |
| Kistler 2021 | ↑ Documentation of surrogate 22.7% → 35.5% ($p = 0.03$);<br>↑ Physician Orders for Life-Sustaining Treatment 9.2% → 18.8% ($p = 0.03$) | | Positive trainee feedback; skills/resources valued |
| Mellinger 2023 | advance care planning discussion/documentation evident in 31% of reviewed charts (trained group) | Not statistically significant |  |
| Mitchell 2018 | No difference in do not hospitalize (adjusted odds ratio 1.08; 95% confidence interval 0.69–1.69) | ↑ No tube-feeding directives at all timepoints (adjusted odds ratio 1.78–2.32);<br>↑ goals-of-care discussion at 3 months (adjusted odds ratio 2.58; 95% confidence interval 1.20–5.54) only<br>no change in burdensome treatments and comfort-care preference unchanged |  |
| Monnet 2024 |  |  | [interactive advance care planning] tools [website] perceived useful to initiate advance care planning; uncertainty about next steps after prompts; need facilitator guidance |
| Moore 2017 |  | Satisfaction with care at End-of-life in dementia: cohort median 30 (interquartile range 29–33); Nursing home 2 median 34 (interquartile range 28–39). |  |
| Sussman 2022 | | ↑ advance care planning engagement—users vs non-users: $4.33 \pm 0.54$ vs $3.40 \pm 0.73$ ($p < 0.01$) | 58% completed workbook; spouse-paired dyads more likely to complete; workbook “non-threatening,” reduces guessing |
| Tilburgs 2020 | ↑ advance care planning discussed/recorded with people with dementia: 49.3% vs 13.9% (intracluster correlation coefficient 0.4, odd ratio 1.99; $p = .002$);<br>↑ advance care planning discussions per patient 2.3 vs 0.2 (adj. $\Delta$ 2.4; 95% confidence interval 1.2–3.5) | No differences in people with dementia’s quality of life, Shared decision-making experience, carer competence, costs, or quality-adjusted life years | |
| Vellani 2023 | | ↑ advance care planning (readiness/self-efficacy): $3.2 \pm 0.6$ → $3.7 \pm 0.8$; 95% confidence interval 0.5 [0.3–0.7]; $p = 0.00014$; $d = 1.1$ | |
| Wils 2017 | Numbers of advance directive ($p = 1.00$) and appointed representatives ($p = 0.08$) unchanged; other advance care planning items ↑ significantly ($p < 0.05$) | ↑ Total advance care planning items in electronic medical record: 39 → 73 at 12 months | |
| Yeung 2023 | | ↑ Self-efficacy at T3 ($p = 0.030$; $d = 0.98$); ↑ readiness at T1 ($p = 0.006$; $d = 1.23$); ↑ concordance at T1–T3 ($\beta$ 2.83–3.64; $p = 0.001$–$0.027$); Zarit burden ↑ at T2 only ($p = 0.040$; $d = 0.56$) | |

## Discussion

This rapid review synthesised evidence from 25 studies (2015–2025) across 12 countries. We have identified three types of advance care planning interventions—using videos as decision aids, web-based tools, and multicomponent programmes—and strategies to discuss advance care planning by addressing families’ lack of knowledge in dementia and using relevant frameworks. In line with the previous reviews (Kelly et al., 2019; McMahan et al., 2021;

Wendrich-van Dael et al., 2020), the synthesised findings indicate that the identified advance care planning interventions prove effective to increase the uptake but their effects on secondary health outcomes (e.g., decisional conflict, concordance, hospitalisation, quality of life) were mixed.

The mixed results could stem from several factors. That is, the included studies variably defining the notion of advance care planning as anything from the completion of an advance directive (Jennings et al., 2019; Kistler et al., 2021) to broader engagement in both medical and non-medical decision-making that are reflective to people with dementia’s values (Bosisio et al., 2021; Dupont et al., 2024; Huang et al., 2022; Huang et al., 2020; Ménard et al., 2025; Monnet et al., 2024; Wils et al., 2017). This variability impedes meaningful cross-study comparisons and the aggregation of evidence across contexts. As a result, the effects of advance care planning interventions resulted in variable secondary health outcomes.

The secondary health outcomes that showed positive effects primarily related to increased advance care planning engagement, attitudes and knowledge of advance care planning, concordance of care, decisional autonomy, family perception of care as well as reduced decisional conflict.

Still, many studies produced mixed results over the effects of advance care planning over people with dementia’s quality of life, their experienced level of shared decision making, carers’ sense of competence, health care costs and people with dementia’s quality-adjusted life year (Tilburgs et al., 2020). The rate of burdensome treatment in people with dementia also seem to be not statistically significant (Mitchell et al., 2018). Finally, the hospitalisation rate of people with dementia also seems to be mixed (Bavelaar et al., 2023; Jennings et al., 2019)

In terms of the quality of the studies, only six (24%) included articles were deemed as high quality whereas 44% and 32% were deemed as medium and poor quality respectively.

For randomised control trials, concerns were noted regarding inadequate description of the randomization procedures, participants selection or allocation concealment processes, raising the likelihood of selection bias. This is important given the validity of the results may depend on whether appropriate statistical adjustments were applied. Moreover, most quasi-experimental studies had small sample sizes and relied on simple statistical methods, contributing to additional limitations. For instance, Cohen et al. (2019) restricted the inclusion of certain populations (e.g., Asian populations) and applying culturally limited statistical adjustments. They categorised race only as ‘White’ vs. ‘Others’ thereby further reducing the generalisability of findings.

For qualitative studies, the omissions of or limited discussion around researcher’s reflexivity and positionalities contribute reduce the transferability of these findings (Forero et al., 2018; Olmos-Vega et al., 2023), signalling the need to be addressed in the future.

There is limited evidence that explicitly discusses how cultural norms, resource availability, and national legal frameworks shape how advance care planning interventions are received. This is important to address given advance care planning, and the associated interventions, must be contextually grounded (Phenwan, Sittiwantana, et al., 2025). As such, it is essential to gain a better understanding around cultures, local legal frameworks and policies and their influence over advance care planning. For instance, countries with collectivist cultures may require culturally tailored approaches, such as adaptations of the Respecting Choices model to address family-centred decision making (Grp et al., 2021). This suggestion contrasts with most of western countries where individuality, self-determination and personal agency are the norm (Cheng et al., 2020; Malhotra et al., 2022).

In summary, this review suggests that advance care planning interventions prove useful to improve its uptake with and for people with dementia with varying health outcomes reported. Still, the varying quality of the included studies makes them less likely to be generalisable beyond their contexts.

### Strengths and limitations

We have included a range of study designs and methods that provided an overview of the field and a rich source of data on advance care planning interventions with and for people with dementia and their effects in their respective contexts.

The study methodology is also rigorous with the dual-screening process for study screening and selection, complemented by regular team discussions to refine both the review process and data interpretation. We have engaged with stakeholders throughout via meetings and email communications, contributing to faster knowledge dissemination with already visible real-world impact.

Still, limitations remain:

First, we only included studies published in English hence there might be missing relevant non-English articles. Second, due to the expedited nature of rapid review, we did not include grey literature. We expedited the data extraction and quality appraisal processes. As such, there might be potential misinterpretation of the data. Still, as recommended by Cochrane, we mitigated this limitation by having secondary reviewers double checking the robustness of these processes given our primary aim was to conduct this review in a timely manner while ensuring the methodological robustness (Garritty et al., 2024; Stevens et al., 2025). Third, due to the vast heterogeneity in quantitative research, a meta-analysis could not be performed. Finally, for the included quantitative and mixed method studies, the publication bias might overall affect the included studies’ conclusions and, consequently, our interpretation. That is, most studies with negative results were often unreported (Bradley et al., 2020). However, to the best of our ability, we were impartial and had interpreted the data as they were presented. One suggestion which goes beyond this review might be to encourage all study findings to be more available, as negative results also yield invaluable insights to the field (Bradley et al., 2020).

### Implications for research

Future research should prioritise the development and adoption of core outcomes set specific to advance care planning in dementia, thereby enabling meta-analyses and benchmarking. This implication is similar to the recommendation from the European Association for Palliative Care which aimed to counterargue Lancet’s scepticism around the usefulness of advance care planning in relation to the changed health outcomes (van der Steen et al., 2025).

Studies of advance care planning interventions in dementia have all originated in high-income countries. This geographic imbalance limits our understanding of how cultural norms and resource constraints shape advance care planning uptake, perpetuating inequities in care and need to be addressed. The global projection predicts that more people with living with dementia will reside in low- and middle-income countries with the largest percentage changes coming from the Middle East and North Africa (Nichols et al., 2022). Future studies should include research partners, research sites from these countries to mitigate this gap for more contextually grounded advance care planning interventions.

### Implications for practice

The following recommendations were proposed as a part of the revised Thai national dementia guideline, which can be applicable to other contexts as well:

-Initiating advance care planning as early as possible. Then, leverage the ‘trigger points’ (medical events or a marked decline in people with dementia’s health and independence) to expand and revise the discussion as necessary.

-always assume that people with dementia has mental capacity to make decisions unless proven otherwise.

-always involve family members in any discussion and decisions (unless explicitly stated otherwise), reflecting the collectivist culture in the country and limited healthcare infrastructures.

-simplify or adjust the language used during conversations as needed to accommodate people with dementia and families

-increasing the numbers of dementia advance care planning’s facilitators via training.

## Conclusions

This rapid review demonstrates that contextually tailored advance care planning interventions consistently increases advance care planning uptake in dementia care although evidence on secondary outcomes remains mixed.

The vacillation in advance care planning definitions and outcome measures calls for the urgent need for a standardized core outcome set and methodological harmonization to support meta-analyses and robustly inform clinical practice.

Future research must extend beyond high-income countries to include low- and middle-income settings and people with advanced dementia, addressing equity by incorporating participatory, culturally adapted designs that resonate with local care structures and contexts.

## Data Availability

All data produced in the present work are contained in the manuscript

## Acknowledgments

We would like to express our thanks to Dawn Adams for her help with the search terms and search process.

Declaration of conflicting interest

The author(s) declared no potential conflicts of interest with respect to the research, authorship, and/or publication of this article.

## Funding statement

The author(s) received no financial support for the research, authorship, and/or publication of this article.

*Ethical approval and informed consent statements Ethical approval*

Ethics approval was not required for this rapid review.

## Informed consent

This study does not involve research with human participants. Therefore, informed consent was not required.

## Consent for publication

Not applicable.

## Data availability statement

This article is a rapid review. The protocol was registered on the PROSPERO database (CRD420251034120) and can be accessed at https://www.crd.york.ac.uk/PROSPERO/view/CRD420251034120

## Supplemental Material

Supplemental material for this article is available online.

## CRediT author statement

**Tharin Phenwan:** Conceptualization, Methodology, Software, Validation, Fornal analysis, Investigation, Resources, Data curation, Writing - Original Draft, Writing - Review & Editing, Visualization, Supervision, Project administration

**Thagoon Kanjanopas:** Conceptualization, Methodology, Validation, Fornal analysis, Investigation, Data curation

**Kanthee Anantapong:** Conceptualization, Methodology, Validation, Fornal analysis, Investigation, Data curation, Writing - Review & Editing

**Supakorn Sripaew:** Conceptualization, Methodology, Validation, Fornal analysis, Investigation, Data curation, Writing - Review & Editing

**Jiroj Phalalert:** Conceptualization, Validation, Fornal analysis, Investigation, Data curation

**Atiqur Rahman:** Validation, Fornal analysis, Writing - Review & Editing

## Notes

### Competing Interest Statement

The authors have declared no competing interest.

### Clinical Protocols

https://www.crd.york.ac.uk/PROSPERO/view/CRD420251034120

